# Impact of Measurement Imprecision on Genetic Association Studies of Cardiac Function

**DOI:** 10.1101/2023.02.16.23286058

**Authors:** Milos Vukadinovic, Gauri Renjith, Victoria Yuan, Alan Kwan, Susan C. Cheng, Debiao Li, Shoa L. Clarke, David Ouyang

## Abstract

**Background:** Recent studies have leveraged quantitative traits from imaging to amplify the power of genome-wide association studies (GWAS) to gain further insights into the biology of diseases and traits. However, measurement imprecision is intrinsic to phenotyping and can impact downstream genetic analyses.

**Methods:** Left ventricular ejection fraction (LVEF), an important but imprecise quantitative imaging measurement, was examined to assess the impact of precision of phenotype measurement on genetic studies. Multiple approaches to obtain LVEF, as well as simulated measurement noise, were evaluated with their impact on downstream genetic analyses.

**Results:** Even within the same population, small changes in the measurement of LVEF drastically impacted downstream genetic analyses. Introducing measurement noise as little as 7.9% can eliminate all significant genetic associations in an GWAS with almost forty thousand individuals. An increase of 1% in mean absolute error (MAE) in LVEF had an equivalent impact on GWAS power as a decrease of 10% in the cohort sample size, suggesting optimizing phenotyping precision is a cost-effective way to improve power of genetic studies.

**Conclusions:** Improving the precision of phenotyping is important for maximizing the yield of genome-wide association studies.

**Clinical perspective:** *What Is New?:*

- Measurement imprecision in cardiac imaging phenotypes can substantially impact downstream genetic association studies, explaining much of the difference in identified genetic variants between echocardiography and cardiac magnetic resonance imaging.
- Using the example of left ventricular ejection fraction as an important but imprecise clinical measurement, the analysis suggests that the measurement variation within the range of clinician interpretation reduced genome-wide association studies’ power to detect genetic risk factors as much as decreasing the study population size by 20%.

*What Are the Clinical Implications?:*

- More precise measurements can result in a better understanding of the genetics of cardiac phenotypes and accelerate the development of precision medicine.
- Rather than simply increasing population size, improving measurement precision allows for cost-effective discovery of genetic associations.

## Introduction

Cardiovascular disease is the leading cause of death in the world, and significant work has been undertaken to understand the mechanisms of disease and develop preventive measures. By studying the human genome, insights have been obtained to understand pathways and mechanisms of function and disease risk, and in recent studies, researchers have moved beyond binary labels of disease diagnosis to quantitative phenotypes to obtain greater power in assessing the relationship between genotype and phenotype^1–4^. From quantitative laboratory biomarkers elucidating the relationship between hypercholesterolemia and coronary artery disease^5^ to imaging characteristics in population cohorts^4^ revealing the genetic determinants of cardiovascular development ^6,7^, quantitative assessments of health provide additional signal compared to conventional binary labels of disease.

Despite its relative frequency, critical public health importance, and often penetrant inheritance, heart failure has relatively few known genetic risk factors. Early classic genetic studies were not able to identify many genetic associations with measurements determined by echocardiography^8^. Different cohorts resulted in heterogenous findings in the genetic determinants of cardiomyopathy. More recent studies with measurements from cardiac magnetic resonance imaging (MRI) have been able to find additional loci of relevance and reaffirm previously suspected variants^8,9^, suggesting both larger sample sizes as well as improvements in phenotyping precision, can improve our understanding of the human disease. However, as populations increase, the scaling of phenotyping becomes more costly, and automated approaches are used for most quantitative measurements, despite only moderate correlation compared to manual measurements (R^2^ = 0.318 for cardiac MRI LVEF)^9^.

While quantitative traits have more power than often binary labels of disease, quantitative traits can have noisy measurements. For example, left ventricular ejection fraction (LVEF) as measured by echocardiography can have measurement variation up to 7 - 10%^10,11^, impacting downstream analyses. While cardiac MRI is frequently thought of as a reference standard, given its improved image quality and high signal-to-noise ratio, automated approaches similarly have measurement variation of between 3 – 5%^9^. We use LVEF, an important metric of cardiac function, as an example of an important but noisy measurement to explore the impact of measurement variability on downstream genetic association studies. We compare various methods to obtain the same phenotypic measurement as well as introduce simulated noise in the phenotype measurement to evaluate the relative impact of measurement imprecision and sample size on downstream genetic studies.

## Methods

### Cohort

The UK Biobank is a population-based cohort that links genetic and phenotypic data for approximately 500,000 adult participants from the United Kingdom ^12,13^. We focused on 39,624 participants who had InlineVF measured LVEF ^14^, cardiac MRI, and genetic data available. Before running Genome-Wide Association Studies this cohort was passed through additional quality check filters (**Supplementary Figure 1)**.

### Multiple Approaches to Measure LVEF

Multiple methods of calculating LVEF from the same underlying imaging data were used to assess the impact of phenotyping precision on downstream analyses. First, the UKB provides automated LVEF measurements derived from MRI using Inline VF software^9^, however, this is presented without manual quality control. To compare alternative automated approaches, we also derived LVEF from MRIs using the deep learning segmentation approach suggested by Bai et al ^10^. From the short-axis view videos, segmentation was performed and we calculated the LV volume for each frame using Simpson’s method.

To simulate reader variability, additional experiments were performed introducing Gaussian noise with a mean of 0 and a standard deviation (sd) ranging from [1,10]. We generated multiple phenotypic measurements from the same underlying imaging data, gradually incrementing Gaussian noise, and performed GWAS on each to investigate how measurement error/imprecision affects genetic associations.

Additionally, we further compared results with two final approaches to assess LVEF. When visually assessing LVEF, clinicians often round the value to the nearest 5%, thus we generated a set of phenotype labels by rounding LVEF values to the nearest multiple of 5. For the final comparison, we generated binary LVEF labels by categorizing values as normal or abnormal, with normal values ranging from 52-72 for males and54-74 for females.

### Genome-wide association study

We used the UKB imputed genotype calls in BGEN v1.2 format. Samples were genotyped using the UK BiLEVE or UK Biobank Axiom arrays. Imputation was performed using the Haplotype Reference Consortium panel and the UK10K+1000 Genomes panel^12^. We used the QC files provided by UKB to create a GWAS cohort consisting of subjects who did not withdraw, were of inferred European ancestry, and were unrelated. Subjects with a genotype call rate < 0.98 were also removed. We considered variants with a minor allele frequency (MAF) ≥ 0.01, and we required genotyped variants to have a call rate ≥ 0.95 and imputed variants to have an INFO score ≥ 0.3. Variants with a Hardy-Weinberg equilibrium P value < 1×10^−20^ were excluded. After variant filtering, we were left with 9774199 filtered variants. GWAS was done on a Spark 3.1.1 cluster, using the library Hail 0.2 with Python version 3.6. The GWAS was adjusted for age at MRI and sex. We used the conventional P value of 5×10^−8^ as the threshold for defining genome-wide significance.

### Assessing Association Power’s Relationship with Cohort Size

Apart from noise in phenotype measurements, we also evaluate the effect of cohort decrease on GWAS results. We generated 6 different phenotype files where, starting from the original LVEF cohort (39,624), we keep 90% (35,661), 80% (31,699), 70% (27,736), 60% (23,774), 50% (19812), and 40% (15850) of the samples. Cohort decrease was performed before GWAS QC, and for each step the selection of samples to be excluded was random. Inspecting the effect of cohort decrease helps us define the relationship between the number of LVEF samples and GWAS power.

### SNP-based accuracy

We use an accuracy metric to determine the amount of overlap in significant SNPs between the baseline GWAS results and noise-modified GWAS results. First, we remove all non-significant SNPs by excluding SNPs with a p-value less than *5 × 10*^−8^, which is the Bonferroni corrected p-value threshold. Then, we consider significant SNPs found in both the base results and noise-modified results as true positives (TP), the SNPs found only in the noise-modified results as false positives (FP), and the SNPs not found in the noise-modified results but found in the base results as false negatives (FN). We then calculate 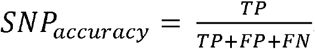.

### GWAS Sensitivity

Sensitivity determines the amount of overlap in significant loci between the baseline GWAS results and noise-modified GWAS results. Specifically, given that *peaks*_*base*_ is the number of significant loci in base GWAS, and *peaks*_*correct*_ is the number of significant loci that persisted in noise GWAS then 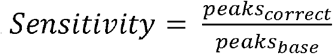. The number of loci and their position can be determined by manual inspection, but we also developed an automatic method. Our automatic method uses a hierarchical clustering algorithm to determine the number and the position of loci from both GWAS, which we then use to compute *peaks*_*base*_ and *peaks*_*correct*_..

## Results

### Quantitative phenotypes improve power of association studies

The study cohort for all analyses consisted of 39,624 adult unrelated subjects of European ancestry (**Table 1**). As a baseline, we first conducted a GWAS of the LVEF phenotype released with the UKBB cardiac MRI data. We identified 5 loci at genome-wide significance on chromosomes 1, 6, 8, 10, and 19 near genes ZBTB17, CDKN1A, CTSB, BAG3, and AP1M1 (**Figure 1)**. In comparison, for an LVEF phenotype binarized to simply abnormal or normal, multiple previously detected loci lost genome-wide significance (including loci for CTSB and AP1M1). Similarly, recognizing the inherent variation present in measuring LVEF, we additionally compared the results if the LVEF was bucketed to 5% bins and showed such imprecision decreased statistical power in all SNPs in the association study compared to the continuous LVEF baseline phenotype.

**Table 1.** Cohort baseline characteristics.

| **Characteristic** | **Mean or n** |
| --- | --- |
| N | 39624 |
| Age at MRI | 54.9 ± 7.47 |
| Male | 18933 (47.8%) |
| Self-identified White British | 33726 (85.1%) |
| Body mass index (kg/m<sup>2</sup>) | 26.5 ± 4.19 |
| Hypertension | 2487 (6.3%) |
| Pulse rate | 67.9 ± 10.9 |
| LV ejection fraction (%) | 55.4 (6.78) |
| LV end diastolic volume (mL) | 141 |
| LV end systolic volume (mL) | 64.1 |

**Figure 1.**
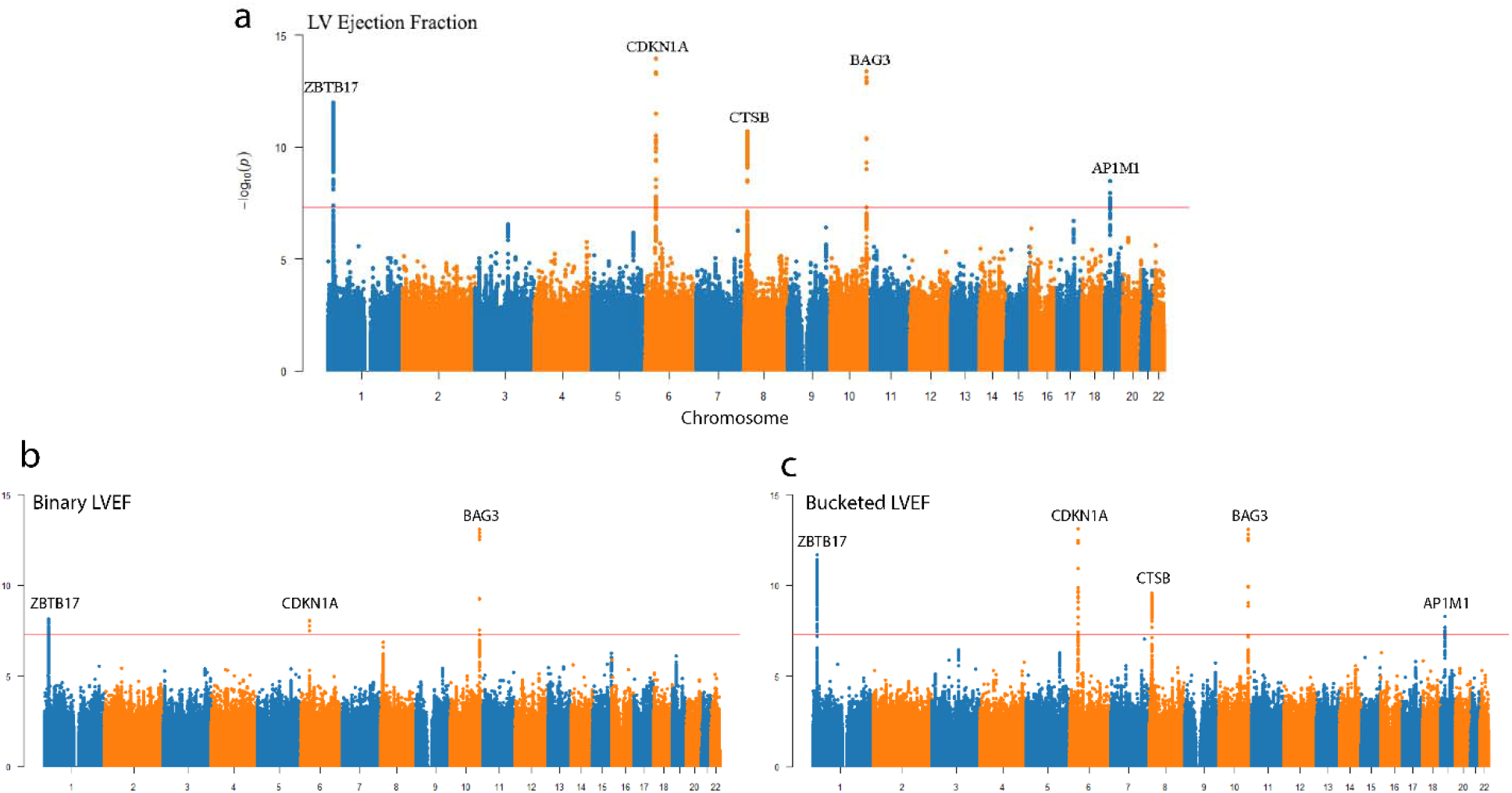
Manhattan plots for genome-wide association studies on UK Biobank reported left ventricular ejection fraction **a**, GWAS on continuous LVEF measurements **b**, GWAS on Normal/Abnormal LVEF where the range for normal is 52-72 in male and 54-74 female population **c**, GWAS on LVEF bucketed to the nearest multiple of 5

### Phenotype noise degrades power of association studies

To investigate the effect of measurement imprecision on GWAS power, we performed a series of association studies while introducing noise in the range of known clinician variation **(Figure 2)**. Simulated variation to the LVEF measurement naturally increases in mean absolute error. Noise with a gaussian standard deviation of 5 results in a mean absolute error of 3.97% and R2 of 0.65 (**Supplementary Table 1)**, and results in the loss of genome-wide significance for the AP1M1 loci on chromosome 19. As we increase phenotypic noise in the range of clinical variation, power gradually declines and the noise equivalent to 7.92% MAE results in a complete loss of genomic-wide significance (**Table 2**). Given echocardiography is known to have a clinician-to-clinician variation of the same or greater MAE^10^, such measurement imprecision could contribute to the limited hits in historial echocardiography-derived GWAS^8^.

**Figure 2.**
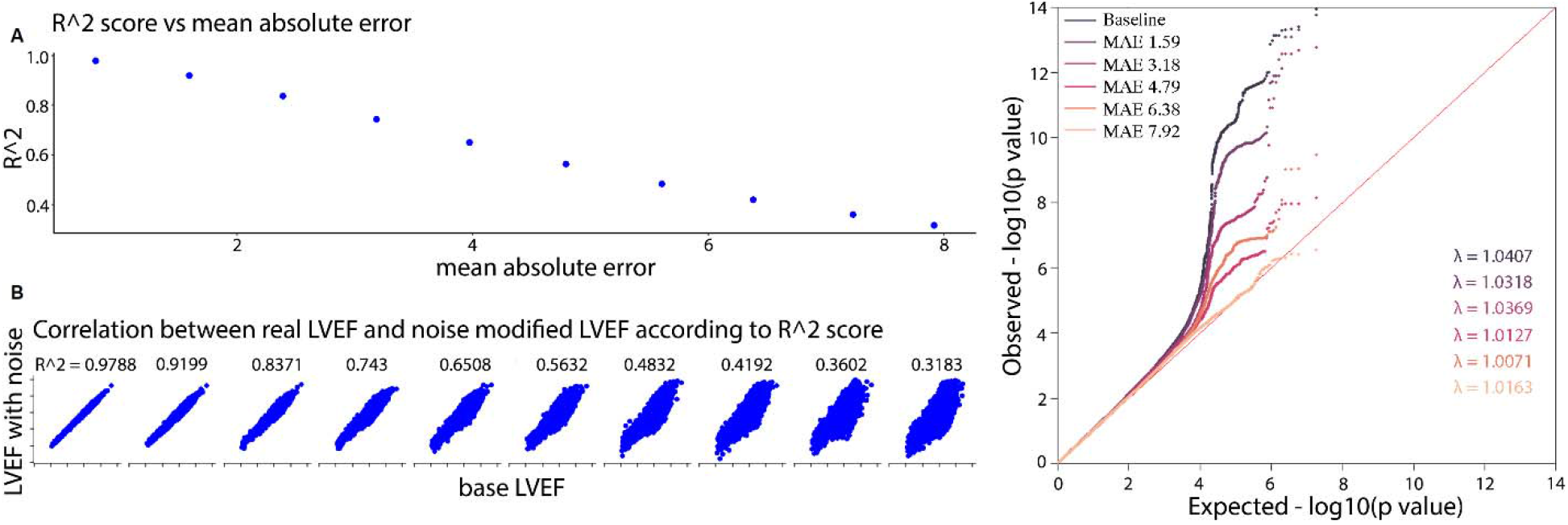
Impact of noise in LVEF on GWAS **a**, Visualizing r2 score, mean absolute error, and the distribution of noise-modified-LVEF with respect to the baseline LVEF **b**, Q-Q plots of P values from GWAS summary statistics for different levels of noise

**Table 2.** Metrics of genetic signal for each increase in SD.

| **Noise SD** | **SNP Accuracy** | **Loci Sensitivity** |
| --- | --- | --- |
| **0%** | 1.0 | 1.0 |
| **1%** | 0.9377 | 1.0 |
| **2%** | 0.8547 | 1.0 |
| **3%** | 0.3675 | 1.0 |
| **4%** | 0.2537 | 0.8 |
| **5%** | 0.3921 | 0.8 |
| **6%** | 0.0228 | 0.4 |
| **7%** | 0.0307 | 0.4 |
| **8%** | 0.0145 | 0.4 |
| **9%** | 0.0020 | 0.2 |
| **10%** | 0 | 0 |

### Comparison of Impact of Phenotype Noise vs Cohort Size

Given the summary statistics from 16 different GWAS, we modeled the relationship between noise and GWAS power (**Figure 3, Figure 4)**. There is a linear relationship between the increase in MAE and the decrease in GWAS power. We calculated that an increase of 1% in MAE causes the loci sensitivity to decrease by 13% (p=5.5e-6) and the SNP accuracy by 14% (p=6.6e-5). Experiments with other methods of introducing noise in assessing LVEF similarly show a decrease in genetic association with more imprecise measurements (**Supplementary Figure 2 and Supplementary Table 2**). A similar effect occurs with reductions in cohort size, as a 1% decrease in cohort size results in a 1.3% decrease in loci sensitivity (p=0.01) and a 1.9% decrease in SNP-based accuracy (p=0.0007). We found that a 1% MAE increase has the same effect on loci sensitivity as a 10.3% cohort decrease and the same effect as a 7.2% cohort decrease on SNP accuracy.

**Figure 3.**
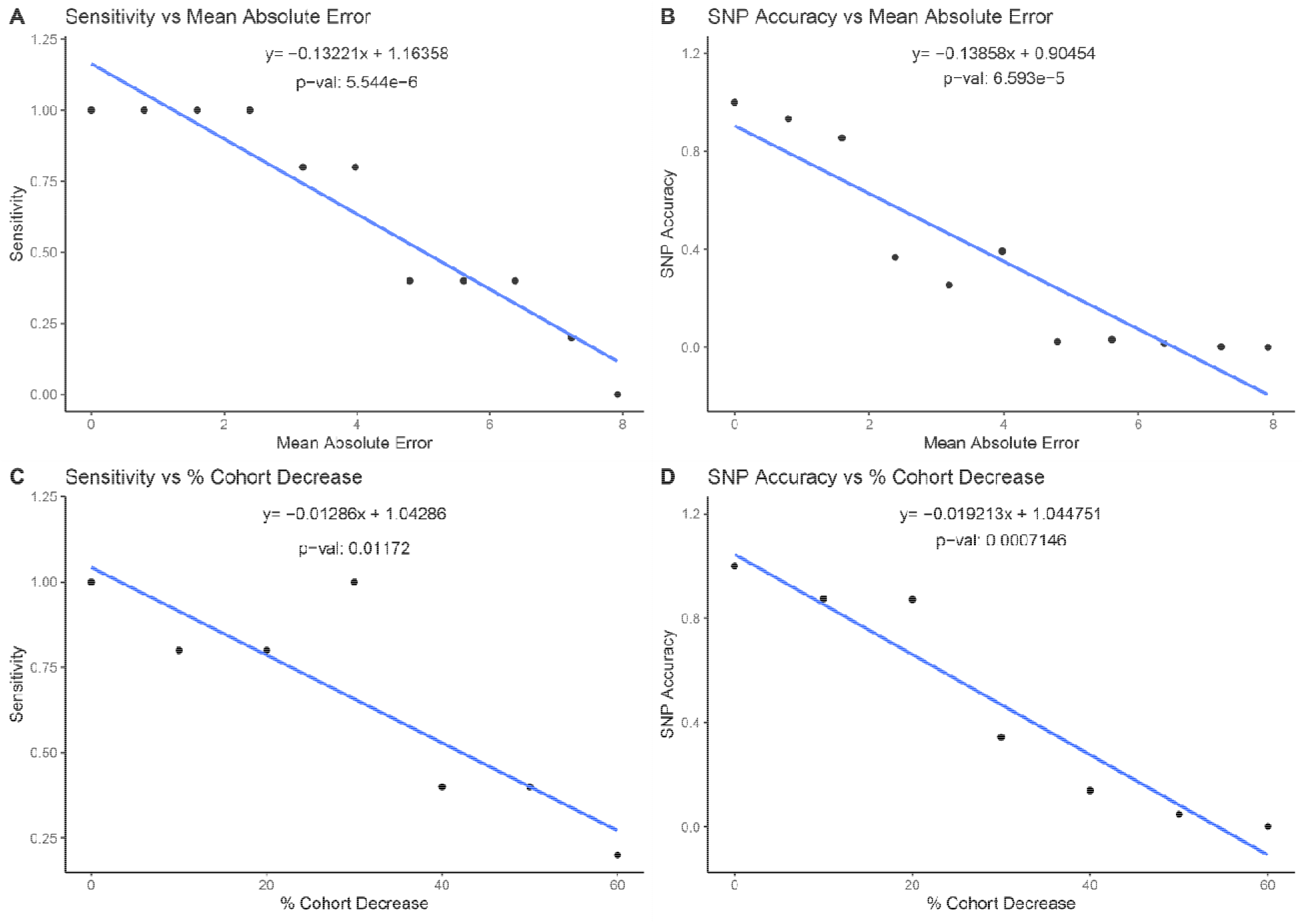
Impact of cohort decrease and noise generation on GWAS power. **a**, Regression analysis on the impact of measurement error quantified by a mean absolute error on sensitivity. **b**, Regression analysis on the impact of the mean absolute error on SNP accuracy. **c**, Regression analysis of the impact of cohort size decline on sensitivity. **d**, Regression analysis of the impact of cohort size decline on SNP accuracy.

**Figure 4.**
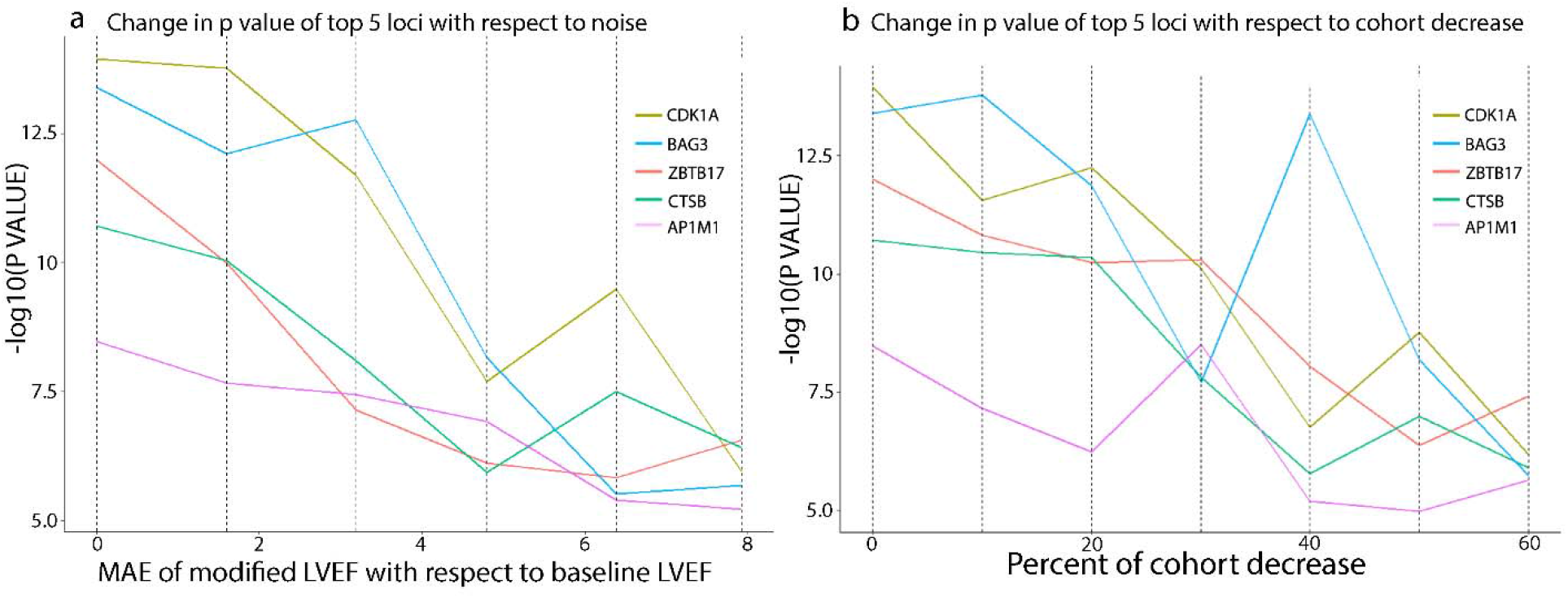
**a**, Slope chart shows the change in the P value of the top 5 loci with respect to mean absolute error; **b**, Slope chart shows the change in a P value of top 5 loci with respect to the cohort decrease; each locus is named after the closest gene

### Improving phenotyping augments downstream genetic analyses

Cardiac MRI provides clinicians and researchers with a plethora of high-resolution imaging, with even the abbreviated 20-min UK Biobank cardiac MRI protocol resulting in 9 sequences with over 30,000 images per study^11^. With so many images and patients, the released UKBB measurements were generated using a fully automated workflow (with Siemens inLineVF) without quality inspection and bias correction. When compared with manual clinician evaluation, the automated measurements of LVEF result in a mean absolute error of 3.4%, R2 of 0.348, and ICC of 0.521 for LVEF. Using a previously published deep learning segmentation model^16^, we independently derived LV segmentation-based calculated LVEF and found a mean absolute error of 6.1%, R2 of 0.335, and ICC of 0.431 for LVEF compared to the automated measurements from UKBB (**Figure 5)**. However, with these deep learning segmentation derived LVEF measurements, the same cohort identified more loci of interest with significant experimental data backing its relevance. In particular, loci on chromosomes 2, 5, and 8 near genes TTN, DNAJC18, and ZNF572 were not previously identified using the released UKBB LVEF measurements but able to be picked up with our quality-controlled measurements.

**Figure 5.**
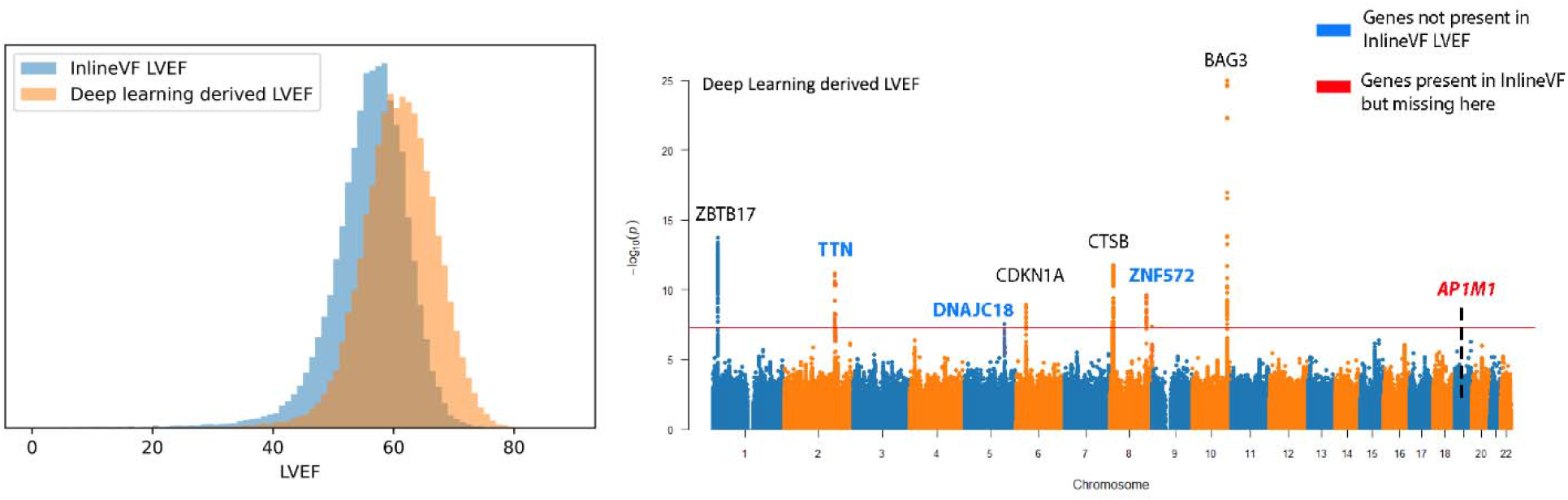
Differences in distribution and GWAS summary statistics between two methods of obtaining LVEF from MRI **a**, Histograms of InlineVF derived LVEF and Deep Learning derived LVEF **b**, Manhattan plot from GWAS performed on Deep Learning derived LVEF; genes colored in blue don’t appear in InlineVF LVEF GWAS (Figure 1a); genes colored in red appear in InlineVF LVEF GWAS but not in deep learning derived LVEF GWAS

## Discussion

In this study, we assessed the impact of measurement imprecision on genetic associations with LVEF and found substantially impaired power in downstream GWAS analysis with even slight increases in imprecision. Even modest phenotyping variation significantly impacted downstream genetic associations, often to a greater extent than changes in cohort size. As measurement variation is present in many clinical measurements and large cohorts increasingly rely on automated measurements, efforts to improve the precision of measurements can potentially be a cost-effective way to maximize the yield of genetic association studies.

Cardiac function as measured by LVEF is an important clinical measurement that defines disease and identifies patients who are eligible for life-prolonging therapeutics. In echocardiography, human test-retest evaluation of LVEF can range between 7-10%, with slight changes in annotation as well as timing that can significantly impact calculations^10,16^. Few variability studies have been undertaken in cardiac MRI, although improved degrees of manual measurement variability have been found^9,17^. Our analysis suggests that a substantial gain in signal comes from the improvement of measurements precision, such that it’s impact is often greater in relative size than sample size. Efforts to improve phenotyping precision that can significantly affect the power and accuracy of downstream analyses.

Noise in measurements can appear in both semi-automated and fully automated workflows^17^, and by improving the precision of measuring LVEF, we also improve the accuracy and robustness of downstream GWAS results. The relatively large improvement in yield of genetic association with more precise phenotyping was substantial in comparison to the marginal benefit of increasing the cohort size. As more genetic analyses are undertaken with automated measurements or assessments^4,7,18,19^, an additional evaluation must be taken to assess the variability and quality of the phenotyping. Such insights ideally will be confirmed with parallel measurements of similar phenotypes or multiple approachs to obtain the same phenotype

In summary, genetic association studies on imaging phenotypes allow researchers to discover many associations that help understand the underlying biology of the disease and structure^20^. For LVEF, even advanced imaging has variability in measurements that can substantially impact downstream association studies. The impact of such variability is even more profound than significant changes in cohort size, suggesting improvement in imaging precision and precise phenotyping in general has significant additional value in improving the power of genetic association studies.

## Data Availability

This paper analyzes UK Biobank data, which is available after the approval of an application at https://www.ukbiobank.ac.uk.

## Acknowledgments

not applicable

## Supplemental Material

### Supplementary Tables

**Supplementary Table 1.** Mapping between Gaussian Noise SD and MAE.

| SD | MAE | R2 |
| --- | --- | --- |
| 0 | 0 | 1 |
| 1 | 0.797489 | 0.9788 |
| 2 | 1.594416 | 0.9199 |
| 3 | 2.386753 | 0.8371 |
| 4 | 3.183924 | 0.743 |
| 5 | 3.974958 | 0.6508 |
| 6 | 4.793956 | 0.5632 |
| 7 | 5.604129 | 0.4832 |
| 8 | 6.380848 | 0.4192 |
| 9 | 7.228321 | 0.3602 |
| 10 | 7.920860 | 0.3183 |

**Supplementary Table 2.**
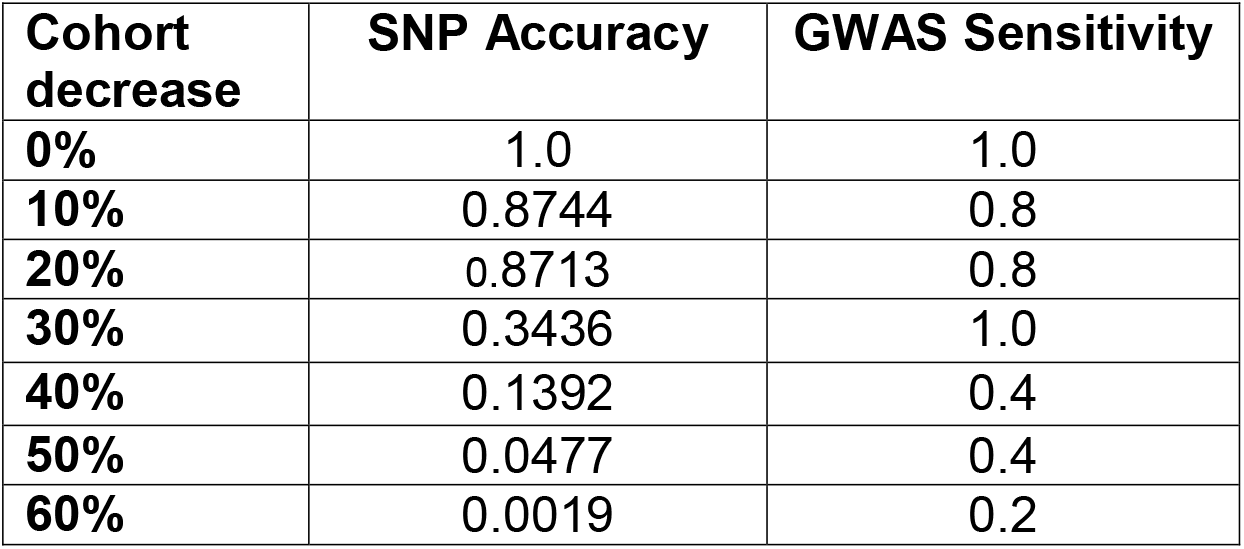
Metrics of genetic signal for each decrease in cohort size.

### Supplementary Figures

**Supplementary Figure 1.**
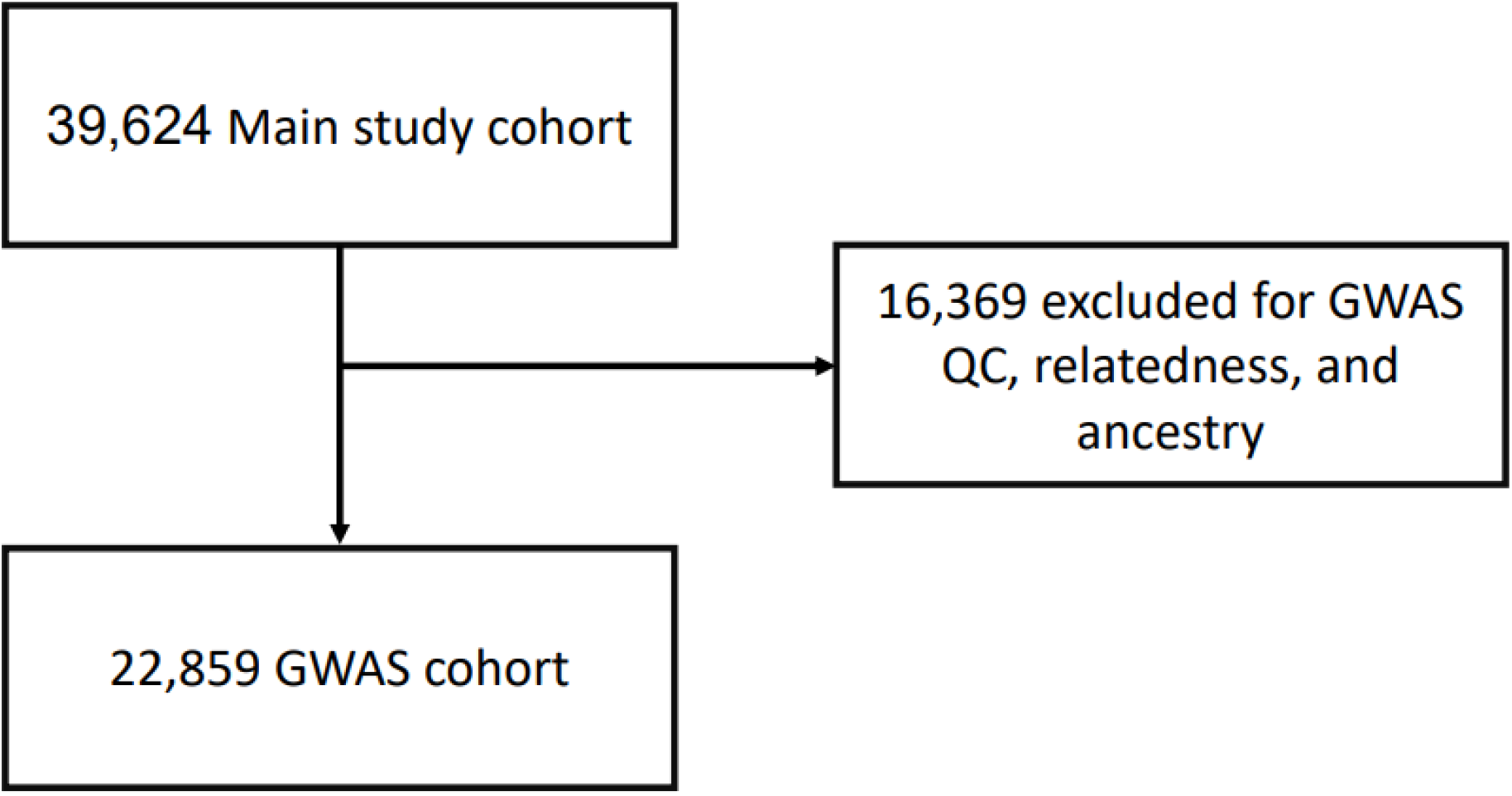
Cohort diagram.

**Supplementary Figure 2.**
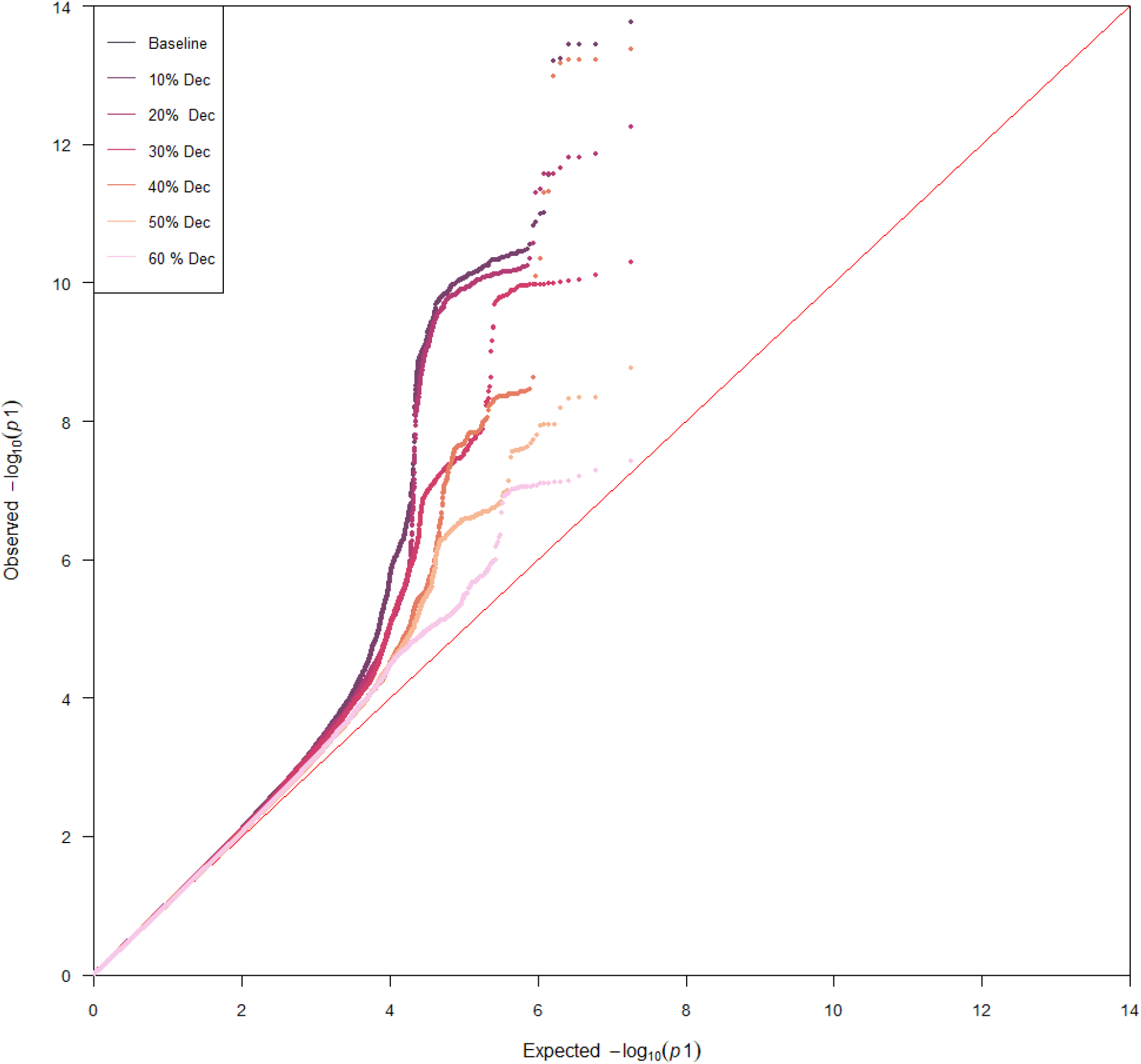
Q-Q plots of P values from GWAS summary statistics for different percentages of cohort decrease

